# Reduced motor unit discharge rates in gastrocnemius lateralis, but not in gastrocnemius medialis or soleus, in runners with Achilles tendinopathy

**DOI:** 10.1101/2022.05.05.22274750

**Authors:** Gabriel L. Fernandes, Lucas B. R. Orssatto, Raphael L. Sakugawa, Gabriel S. Trajano

## Abstract

**Objectives:** Deficits in muscle performance could be a consequence of a reduced ability of a motor neuron to increase the rate in which it discharges. This study aimed to investigate motor unit (MU) discharge properties of each Triceps surae muscle (TS), and TS torque steadiness during submaximal intensities in runners with Achilles tendinopathy (AT).

**Methods:** We recruited runners with (n=12) and without (n=13) mid-portion AT. MU discharge rate was analysed for each of the TS muscles, using High-Density surface electromyography during 10 and 20% isometric plantar flexor contractions.

**Results:** MU mean discharge rate was lower in the Gastrocnemius lateralis (GL) in AT compared to controls. In AT, GL MU mean discharge rate did not increase as torque increased from 10% peak torque, 8.24pps (95%CI: 7.08 to 9.41) to 20%, 8.52pps (7.41 to 9.63, p=0.540); however, in controls, MU discharge rate increased as torque increased from 10%, 8.39pps (7.25 to 9.53) to 20%, 10.07pps (8.89 to 11.25, p<0.001). There were no between-group difference in Gastrocnemius medialis (GM) or Soleus (SOL) MU discharge rates. We found no between-groups differences in coefficient of variation of MU discharge rate in any of the TS muscles nor in TS torque steadiness.

**Conclusion:** Our data demonstrates that runners with AT may have a reduced neural drive to GL, failing to increase MU discharge rate to adjust for the increase in torque demand. Further research is needed to understand how interventions focusing on increasing neural drive to GL would affect muscle function in runners with AT.

## 1. Introduction

Achilles tendinopathy (AT) is one of the most common running injuries, ranging from about 6.2% to 9.5% of all running injuries^1,2^. AT is an overloading injury and although its aetiology is multifactorial^3^, deficits in muscle performance is suggested to be a key factor^4,5^, which seems to be maintained long after symptomatic recovery^6^. Several lines of evidence suggest that neural changes to the triceps surae might underpin some of these chronic motor deficits^7,8^. In particular, it has been shown that individuals with AT have: 1) reduced contribution of gastrocnemius lateralis (GL) to produce plantar flexor force^8^ and b) greater levels of intra-cortical inhibition associated with reduced plantar flexor endurance during single leg heel raise test^7^ when compared to controls. Collectively, these findings suggest that changes in how the central nervous system control specific muscles within the triceps surae might impact load distribution to the tendon in individuals with AT. This is of particular importance because altered triceps surae recruitment due to reduced individual muscle contribution to muscle force could create uneven loading of the Achilles tendon and contribute to tendinopathy^9^.

There has been some speculation about differences in recruitment strategies within the triceps surae in people with AT, with conflicting evidence about which muscle is affected. One study^4^ suggested soleus (SOL) would be the main muscle responsible for the strength and endurance deficits observed in these individuals. The AT group had deficits in plantar flexor torque during dynamometry testing, irrespectively of knee position (knee flexed/extended) compared to controls. The authors reasoned that if the gastrocnemii were affected, deficits between groups would be larger during knee extended and smaller during knee flexed. However, determining force deficits in SOL in relation to the gastrocnemii solely by comparing torque measures between flexed and extended knee positions provides very limited and possibly inaccurate information about muscle recruitment patterns, neglecting all the neurophysiological mechanisms that enable force production in the first place^10^. Conversely, runners with acute AT^8^ (< 3 months) have about 22% lower contribution of GL during 20% and 40% of peak plantar flexor isometric torque but no differences in gastrocnemius medialis (GM) or SOL, compared to controls. Force-sharing contribution of individual muscles of the triceps surae was estimated for each muscle based on the root mean squared (RMS) of surface EMG (electromyography) signal amplitude. Even though data from surface EMG signal is somewhat limited in estimating changes in neural drive to a specific muscle^11^, this result suggests that individual muscles recruitment strategies might be altered in AT.

From a neurophysiological perspective, the force exerted by a muscle depends, partly, on the recruitment and discharge rates of the motor units^10^. Thus, deficits in motor performance could be a result of a reduced ability to recruit motor units and/or to increase the rate at which motor neurones discharge^10^. The analysis of individual motor unit discharge rates from each muscle of the triceps surae ^12^ is a more accurate and robust way of investigating the central nervous system strategy of recruitment of the triceps surae muscle (i.e. neural drive), compared to the typical and limited interference EMG ^13^. This method has been also used in other studies to estimate changes in neural drive to specific muscles in individuals with ACL injury^14^ and patellofemoral pain^15^. Thus, if individuals with AT have reduced neural drive to one or more muscles of the triceps surae, motor unit mean discharge rate of the affected muscle would be reduced compared to the other muscles and to controls^8,16,17^.

Furthermore, reduced control of the plantar flexors could create tendon overload, progressing to early stages of tendinopathy^9^. Increased fluctuation in torque (torque steadiness) is associated with painful musculoskeletal conditions such as knee osteoarthritis or patellofemoral pain^15,18^, and could occur as consequence of greater variation in motor unit discharge rate ^19^. Coefficient of variation of motor unit discharge represents, at an individual muscle level, the ability to effectively control muscle torque and it is an important measure that can help explain motor performance^10^. Chronic musculoskeletal disorders display reduced torque steadiness, such as individuals following ACL reconstruction^20^ or patellofemoral pain^15^. Reduced control of the plantar flexors could lead to tendon overload, progressing to early stages of tendinopathy^9^.

This study aimed to: i) investigate differences in neural drive to each muscle of the triceps surae during submaximal plantar flexor contractions in individuals with AT; ii) determine between groups differences in coefficient of variation of motor unit discharge rate and torque steadiness. We hypothesised that motor unit mean discharge rate of each individual muscles of the triceps surae would be reduced in the tendinopathy group, associated with differences between muscles of the triceps surae in the AT group. We also hypothesised the AT group would have increased variability in motor unit discharge rate and torque steadiness compared to controls.

## 2. Methods

This was a cross-sectional study comparing runners with and without mid-portion AT. A Sample size of 18 participants (9 per group) was calculated based on a similar study^17^ (GPower software parameters: effect size F=0.40; α err prob: 0.05; power 0.95; n=9 per group). 25 endurance runners were recruited for this study, 12 with mid-portion AT (6 males, 44.3 years old **±** 95% CI **7**.6, 173 cm ± 5.7, 76.2 kg ±9.3) and 13 healthy controls (7 males, 34.0 years old ± 4.0, 171 cm ± 5.2, 64.8 kg ±7.1), with a running routine of more than twice weekly for more than 4 months. Runners were recruited from local running clubs, via email and social media. Participants characteristics and torque measures: plantar flexor peak isometric torque and explosive torque, which were not different between groups, have been already partially reported in another study^7^.

All volunteers were endurance runners, recruited from local running clubs around Southeast Queensland, Australia. Diagnosis of mid-portion AT was confirmed by an experienced physiotherapist during examination if patients presented with localised midportion Achilles tendon pain for more than three months, pain provoked by physical activities in a dose dependent way and had pain with palpation at the mid portion of tendon. Volunteers were excluded if presenting insertional AT; previous rupture or surgery of the Achilles tendon; clinical findings indicating a differential diagnosis for the Achilles tendon pain (such as tendon tear); regular participation in other sports involving high speed running (football, rugby, AFL etc), 4) VISA-A score > 90 points for AT group and < 100 for the healthy group; any other musculoskeletal injuries of the lower limb; any neurological disorder; or mental health issues affecting consent. All participants were free of comorbidities such as cardiac, pulmonary, renal, endocrine of gastrointestinal and were not taking any medication for tendon pain or that would affect tendon structure ^21^.

Prior to testing, all participants read and signed a detailed informed consent document and completed the VISA-A questionnaire^22^. The average VISA-A score for the AT was 70.1(± 6.4) and 100 (**±**0) for the control group. The AT group had a running routine of 37.4 (±10.2) km/week and 30.4 (**±**8.4) km/week for the control group. This study was approved by the Queensland University of Technology Human Research and Ethics Committee in line with the Declaration of Helsinki. Data collection was conducted during the COVID-19 pandemic and all safety procedures followed local state government policies.

### 2.1 Data collection and analysis

Plantar flexor isometric peak torque was measured using an isokinetic dynamometer (Biodex Medical Systems, Shirley, New York). For the bilateral AT presentations (n=3), the most symptomatic leg was used and for the control group, the dominant leg was used for testing. Leg dominance was selected by asking the participants what their preferred leg was, and if they were unsure, they were asked which leg they would use to kick a ball. Participants were sitting (75 degrees of hip flexion) with their knee straight and with the foot perpendicular to shank. Warm up consisted of 2 × 4 s isometric contractions of each participant’s perceived 20, 40, 60 and 80% maximal voluntary isometric contraction intensity. After warm-up, participants performed at least 3 maximal voluntary isometric contractions, until <5% variation was observed between contractions, and the highest value was used. Thereafter, participants performed three trapezoidal submaximal isometric contractions (3×10% and 3×20% peak torque) in a randomised order. For each intensity participants had 4 attempts to get familiarised with task before recordings. Rate of torque rise and decline was standardised at 10% peak torque/s between contractions with different intensities, with a 10-s sustained plateau at the top, followed by 1 min of rest between contractions ^23,24^. Participants received real-time visual feedback of the trapezoidal pathway, displayed in a monitor placed at 1m away from the participant. During the plantar-flexors trapezoidal contractions, HD-EMG (Sessantaquattro, OTBioelettronica, Torino, Italy) signals were recorded with OT Biolab+ software (version 1.3.0., OTBioelettronica, Torino, Italy), from SOL, GM and GL. After skin preparation (shaving, light abrasion, and cleansing of area with alcohol), electrodes were positioned following the estimated muscle fibres orientation using a bi-adhesive layer with a conductive paste to ensure good skin-electrode contact and conductibility. One 32-channels electrode matrice (ELSCH032NM6, OTBioelettronica, Torino, Italy) was placed on GM, one 32-channels electrode matrice on GL and two 32-channels electrodes matrices on SOL, one laterally and one medially to the Achilles tendon, to ensure sufficient data were gathered for motor unit analysis. The ground strap electrode (WS2, OTBioelettronica, Torino, Italy) was dampened and positioned around the ankle joint of the tested leg. The EMG signals were recorded in monopolar mode, amplified (256x), band-passed filtered (10-500Hz) and converted to digital signal at 2048Hz by a 16-bit wireless amplifier (Sessantaquattro, OTBioelettronica, Torino, Italy), before being stored for offline analysis. Since the matrice adapter device (AD2×32SE, OTBioelettronica, Torino, Italy) has only 2 channels for electrode connection, each intensity of the protocol had to be performed twice, once with electrodes connected to the gastrocnemii and a second time with the electrodes connected SOL, this order was randomised for each intensity. Torque signal was recorded and analysed with OT Biolab+ software. HD-EMG signal was recorded and analysed offline, decomposed into motor unit spike trains, then converted into instantaneous discharge rates with specialised software using blind source separation decomposition technique using DEMUSE tool software (v.4.1; The University of Maribor, Slovenia) ^25^. For each muscle and for each intensity, the 2 best contractions, with the lowest deviation from trapezoidal torque trajectory, were combined in one file and motor unit tracked across the 2 contractions at the same intensity for analysis. All motor units were visually inspected, erroneous discharge times were excluded, and missed discharges included. Manual inspection is required to reduce automatic decomposition discharge errors and improve data reliability^26^. Only motor units with a pulse-to-noise ratio >30dB, sensitivity > 90%, were used for data analysis ^24,25^. For participants that yield no good quality motor units after motor unit tracking across the 2 contractions at the same intensity, the best single contraction was used for analysis with the motor unit discharge characteristics inspected as mentioned above. Due to the reduced number of the same motor units found across intensities, motor unit tracking across intensities was not feasible and therefore not used for analysis. Motor units were collected from the 10 seconds isometric plateau, the first and last two seconds were excluded and analysis of mean motor unit mean discharge rate, coefficient of variation of motor unit discharge rate and torque steadiness were performed from the central 6 seconds of the isometric plateau. Motor unit mean discharge rate, coefficient of variation of motor unit discharge rate and were calculated for each muscle during 10% and 20% peak torque, trapezoidal contractions.

Torque steadiness was analysed as the coefficient of variation in torque for each torque intensity tested. Torque was filtered (10Hz, 4th order, low pass). To reduce intrasubject variability, data from the coefficient of variability of torque was averaged across the 4 contractions for each of the two torque intensities tested, the 2 contractions recorded during gastrocnemii testing and from the 2 contractions during SOL testing^27^.

### 2.2 Statistical analysis

All analyses were performed using R studio (version 1.3.1093). Models were fitted using the *lme4* package 30. Separate linear mixed-effect models were used to compare motor unit mean discharge rates and coefficient of variation of motor unit discharge rate of identified motor units for each muscle (SOL, GM, GL); between intensities (10% and 20%) and groups (AT and control). We tested the model using a random intercept (participant ID) and slope (recruitment threshold by intensity) for each participant in the study to account for the influence of motor unit populations and the correlation between repeated observations in each participant. The estimated marginal mean difference and 95% confidence intervals (CI) for all variables (motor unit mean discharge rate, coefficient of variation of motor unit discharge rate, between groups and torque steadiness) were determined using the *emmeans* package31. Normality assumptions were confirmed by analysis of the histogram of residuals, Q-Q Plot and the residual-predicted scatterplot. Independent t-test was used to compare torque steadiness between groups for each torque intensity. An alpha level of 5% was set for statistical significance for all tests, and when appropriate, Bonferroni post-hoc analysis was performed. Data is presented as mean (± 95% CI).

## 3. Results

### 3.1 Motor unit identification

We found a total of 1.057 motor units, 518 motor units in the AT (average of 43 per participant) and 539 motor units in the control group (average of 41 per participant) across all muscles and intensities. A total of 120 motor units were found in SOL at 10% peak torque in the AT group and 120 motor units at 20%. In the control group, we found a total of 127 motor units at 10% and 114 motor units at 20% peak torque. GM analysis yield 101 motor units in the AT at 10% peak torque and 128 motor units at 20%. In the control group, we found a total of 113 motor units at 10% and 116 motor unit at 20%. For GL analysis, we identified a total of 20 motor units at 10% peak torque in the AT and 29 motor units at 20%. In the control group, we found a total of 43 motor units at 10% and 26 motor units at 20%. GL was the muscle with the least amount motor unit found in single contractions, and some were lost during motor unit tracking between the two contractions of the same intensity.

### 3.2 Motor Unit discharge rate

Analysis of SOL motor unit mean discharge rate showed difference between torque intensities (F=20.118, p<0.001, η2p=0.04) but no differences between groups (F=0.324, p=0.574) or intensity × group interaction (F=0.512, p=0.474). There was an increase in motor unit mean discharge rate in both groups as torque increased. In the AT, motor unit mean discharge rate increased from 6.98pps (6.72 to 7.24) at 10% peak torque to 7.29pps (7.02 to 7.55) at 20%; the motor unit mean discharge rate in the control group also increased, from 7.40pps (7.16 to 7.63) at 10% peak torque to 7.86 pps (7.55 to 8.16), (Figure 1).

**Figure 1.**
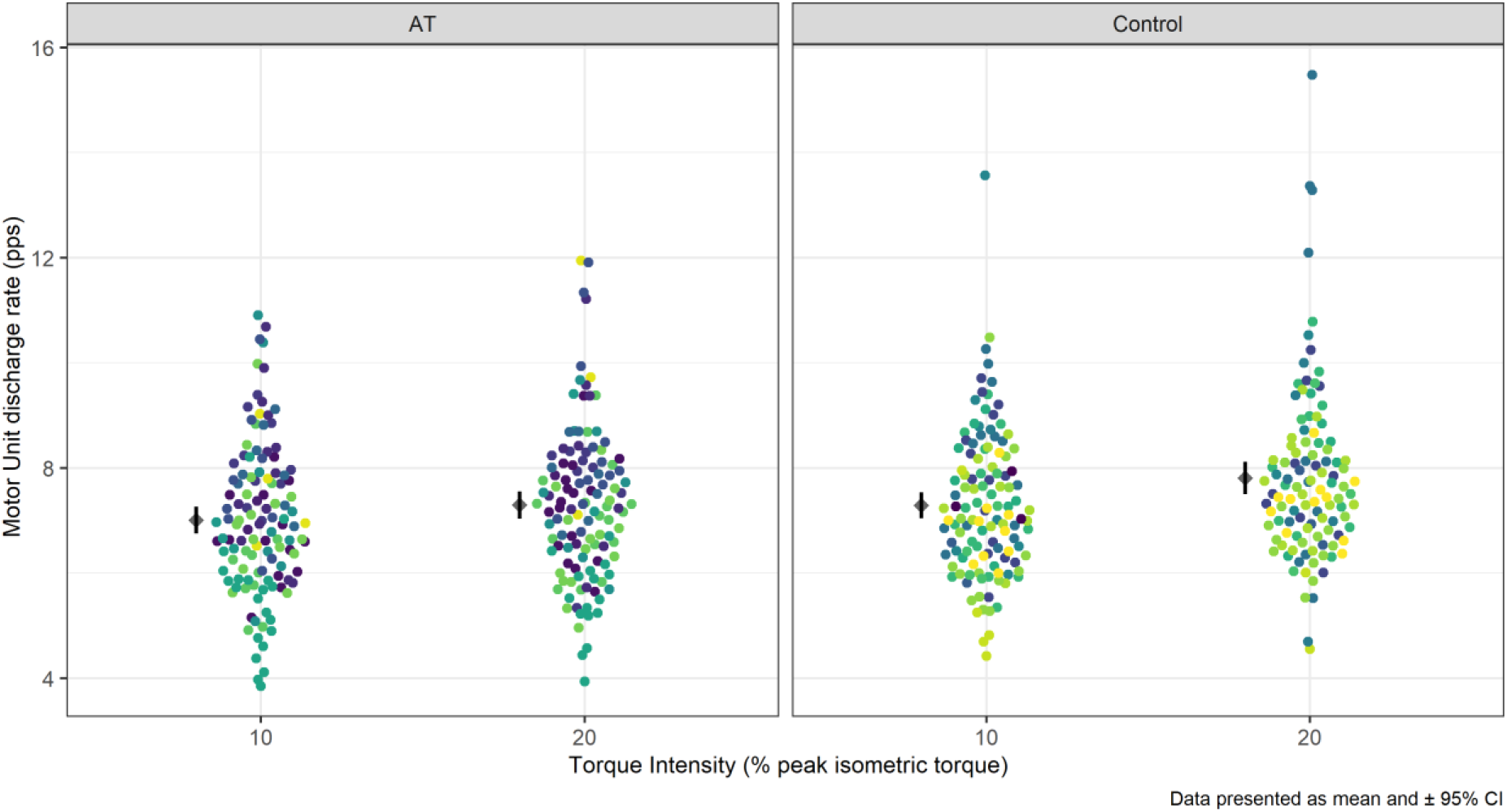
Motor unit mean discharge rate of Soleus during 10% and 20% peak isometric contraction. Each dot represents a single motor unit data point, coloured by participants. Mean and 95% confidence interval are offset to the left to facilitate visualisation. pps = pulse per second.

Similar to SOL, in GM analysis, we observed differences in motor unit mean discharge rate between different torque intensities (F=75.554, p<0.001, η2p=0.15) but no differences between groups (F=0.488, p=0.492) or intensity × group interaction (F=1.063, p=0.303). In both groups, motor unit mean discharge rate increases with the increase in torque intensity. In the AT group, motor unit mean discharge rate increased as torque increased from 10%, 8.38pps (7.44 to 9.33) to 20% peak torque, 9.54pps (8.61 to 10.48). The same was observed in the control group, motor unit mean discharge rate increased as torque increased from 10%, 8.63pps (7.75 to 9.51) to 20%, 10.10pps (9.21 to 11.00) (Figure 2). On the other hand, in GL, we found an intensity × group interaction (F=27.955, p=0.001, η2p=0.11). While in the AT, motor unit mean discharge rate did not change as torque increased from 10% peak torque, 8.24pps (7.08 to 9.41) to 20%, 8.52pps (7.41 to 9.63, p=0.540); however, in the control group, motor unit mean discharge rate increased as torque increased from 10%, 8.39pps (7.25 to 9.53) to 20% peak torque, 10.07pps (8.89 to 11.25, p<0.001), (Figure 3, ** denotes statistical difference). The control group had a higher motor unit mean discharge rate at 20% torque compared to the AT group.

**Figure 2.**
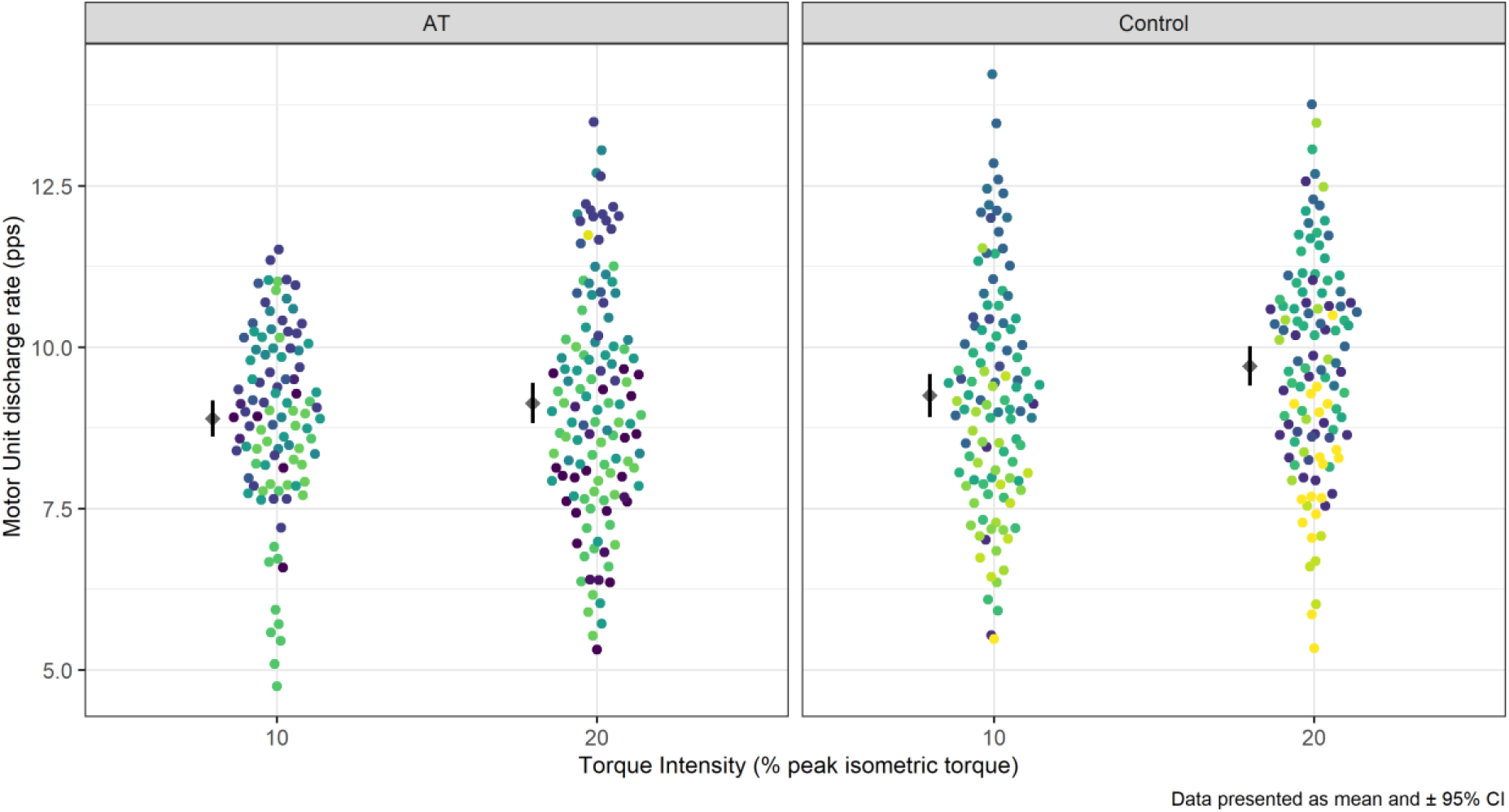
Motor unit mean discharge rate of Gastrocnemius medialis during 10% and 20% peak isometric contraction. Each dot represents a single motor unit data point, coloured by participants. Mean and 95% confidence interval are offset to the left to facilitate visualisation. pps = pulse per second.

**Figure 3.**
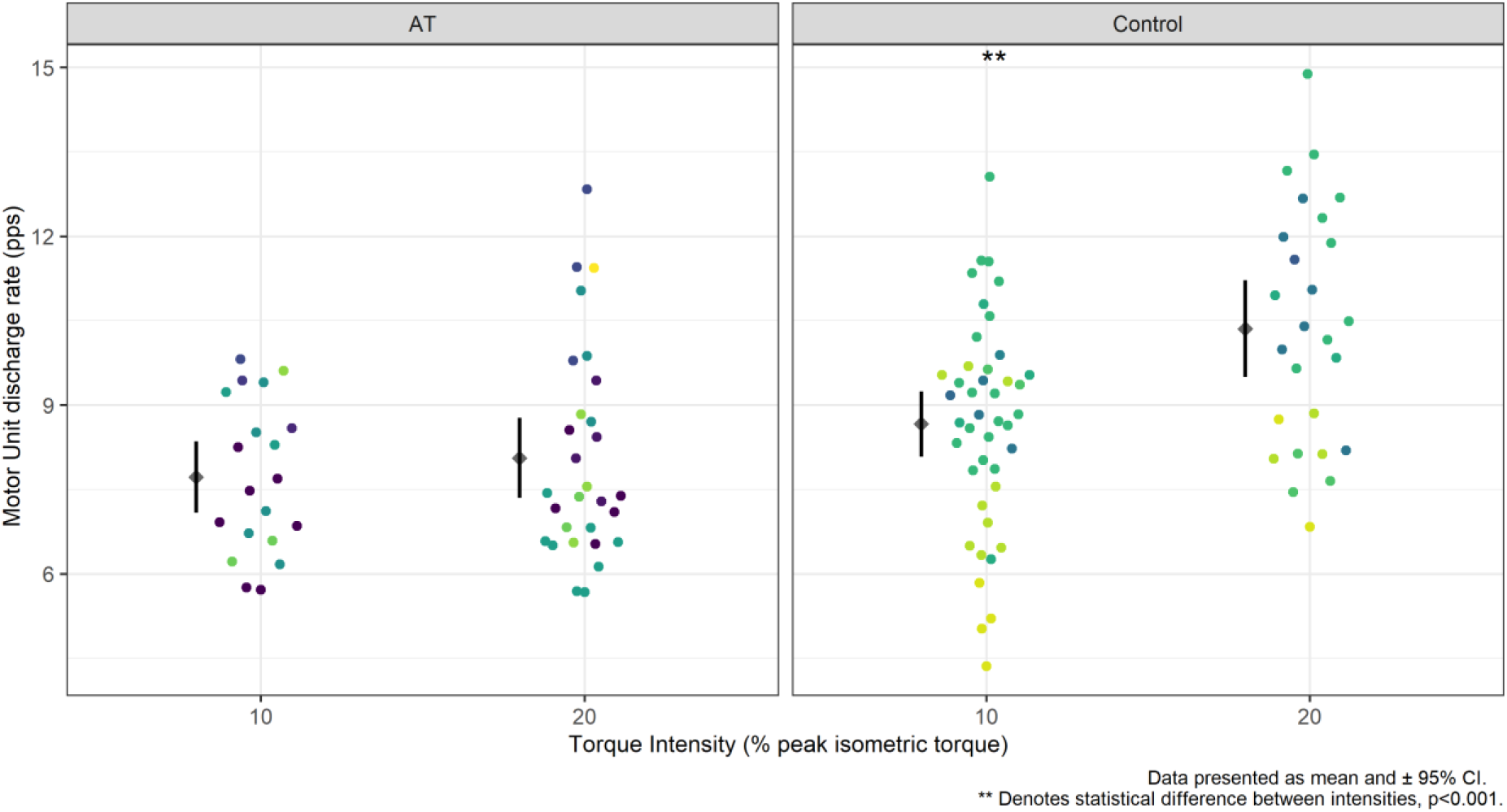
Motor unit mean discharge rate of Gastrocnemius lateralis during 10% and 20% peak isometric contraction. Each dot represents a single motor unit data point, coloured by participants. Mean and 95% confidence interval are offset to the left, to facilitate visualisation. pps = pulse per second. ** denotes statistical difference, p<0.001.

### 3.3 Coefficient of variation of motor unit discharge rate

SOL had no difference in coefficient of variation of motor unit discharge rate between intensities (F=2.963, p=0.086) or between groups (F=0.151, p=0.700). In the AT, the coefficient of variation of motor unit discharge rate was 10.3% (8.8 to 11.8) at 10% peak torque and 9.3% (7.9 to 10.7) at 20%; in the control group, the coefficient of variation of motor unit discharge rate was 10.2% (8.8 to 11.7) at 10% peak torque and 10.0% (8.6 to 11.5). GM presented difference in coefficient of variation of motor unit discharge rate between intensities (F=51.203, p<0.001, η^2^p=0.10) but not between groups (F=3.673, p=0.07) and had no intensity × group interaction (F=0.872, p=0.350). In the AT, the coefficient of variation of motor unit discharge rate was 12.15% (11.1 to 13.1) at 10% peak torque and 9.16% (8.1 to 10.1) at 20%; in the control group, the coefficient of variation of motor unit discharge rate was 10.75% (9.8 to 11.7) at 10% peak torque and 8.45% (7.4 to 9.5). In GL, as well as in GM, we observed difference between intensities (F=5.222, p=0.024, η^2^p=0.05) but not between groups (F=0.661, p=0.428) or intensity × group interaction (F=2.779, p=0.098). In the AT, the coefficient of variation of motor unit discharge rate was 10.9% (8.5 to 13.2) at 10% peak torque and 11.9% (9.6 to 14.0) at 20%; in the control group, the coefficient of variation of motor unit discharge rate was 12.7% (10.5 to 14.9) at 10% peak torque and 12.9% (10.5 to 15.2).

### 3.4 Torque steadiness

There were no differences in torque steadiness analysis between groups in either of the two intensities analysed. Mean coefficient of variation of torque at 10% peak torque, in the AT group was 1.06 (0.79 to 1.32) and 1.13 (0.90 to 1.37, p=0.656) in the control group; and at 20%, mean coefficient of variation in torque in the AT group was 0.80 (0.57 to 1.03) and 0.92 (0.73 to 1.11, p=0.375) in the control group.

## 4. Discussion

### 4.1. Main findings

The present study aimed to determine if runners with chronic mid-portion AT had reduced neural drive to the triceps surae and if there were muscle-specific differences in motor unit discharge characteristics within the triceps surae. For that, we analysed motor unit mean discharge rate and coefficient of variation of motor unit discharge rate of each individual muscle of the triceps surae during isometric contractions of increasing intensities. We also aimed to determine if the AT group had reduced torque steadiness.

Our data indicate that runners with AT have reduced neural drive to GL during the increase in plantar flexor isometric torque output. We confirmed our primary hypothesis, demonstrating a muscle-specific difference in neural drive in the AT group and a reduced neural drive to GL during the increase in plantar flexor torque, not observed in the control group. However, we had also hypothesised that the neural drive to the triceps surae of the AT group would be reduced, but GM and SOL were no different from controls. Furthermore, we did not confirm our second hypothesis, as we found no differences in the coefficient of variation of motor unit discharge rate in any of the muscles, nor did we find differences in triceps surae torque steadiness between groups.

### 4.2. Mean motor unit discharge rates

It has been previously identified that the three muscles of the triceps surae, although synergists as ankle plantar flexors, may have an independent neural drive from one another, allowing independent recruitment strategies for better joint control^12^. Our study also observed independent neural drive within the triceps surae. Further, we found different neural strategies between groups in only one of the three muscles of the triceps surae. Our data show that the AT group does not use the GL as effectively as healthy controls to match the increase in plantar flexor torque intensity. Although there was no difference in motor unit mean discharge rate between groups in GL when comparing the same level of torque, the AT group had a reduced motor unit mean discharge rate with the increase in torque, outlining a change in recruitment strategy in GL that was not observed in the control group. Similar findings have been reported in another study with runners with AT^8^. They used the physiological cross-sectional area and normalised RMS EMG to calculate the index of force of each muscle and estimate individual muscles’ contribution to triceps surae force production. GL had a significantly lower contribution to overall triceps surae force output; thus, reduced neural drive compared to healthy counterparts. Muscle force depends on motor unit discharge rate, which is proportional to the neural drive to the muscle. In healthy individuals, motor unit discharge rate increases to adjust for an increase in torque intensity^10^. Contrary to what was previously suggested in the literature^4^, we found no differences in SOL motor unit mean discharge during the increase in torque, which suggests that SOL contribution to plantar flexor force is not impaired in AT.

The reduced neural drive to GL observed in our study seems relevant in the persistent muscle deficits observed in AT^6^. Perhaps current treatment strategies for AT fail in effectively rehabilitating GL function; therefore, maintaining this reduced neural drive and contribution to force production during ankle plantar flexion. Individual muscles of the triceps surae have independent neural drive^12^. Therefore, finding strategies to increase GL recruitment and contribution during exercise is important, as altered muscle coordination (i.e. individual muscle contribution for force production within a muscle group) may lead to unequal loading to the Achilles tendon^28^. Performing heel raises with the foot positioned with toes pointed inwards, significantly increased GL motor unit discharge rate compared to toes neutral in healthy individuals^12^. Foot position, in healthy individuals, can also selectively affect GM and GL hypertrophy ^29^. Therefore, implementing different foot positions during rehabilitation could help increase GL activity during plantar flexor resistance training. Rehabilitation programs using different foot positions during triceps surae resistance training should be studied in patients with AT to explore how this reduced contribution of GL in triceps surae torque, impacts AT and if implementing treatment strategies to increase the neural drive to GL would influence tendon pain and function in AT.

### 4.3. Coefficient of variation of motor unit discharge rate and Torque steadiness

Torque variability was measured during 10 and 20% relative peak isometric torque plateau. We found no differences between groups in coefficient of variation of motor unit discharge in any of the muscles of the triceps surae nor did we find differences in triceps surae torque steadiness. All three muscles of the triceps surae were equally matched to controls in the two submaximal intensities tested. coefficient of variation of motor unit discharge represents, at an individual muscle level, the ability to effectively control muscle torque and it is an important measures that can help explain motor performance^19,30^. Fluctuations in torque, coefficient of variation of motor unit discharge rate and torque steadiness are more variable in lower intensities than in higher torque intensities, hence why 10 and 20% intensities were used for analysis^19^.

Based on our findings, the ability of the triceps surae in controlling torque during submaximal contractions is not affected in runners with AT, which aligns with another study^31^. Torque steadiness is affected by pain^18^, and reduced torque steadiness has been reported in other chronic^18^ and painful musculoskeletal conditions^15,20^. In our study, we used submaximal torque intensities and none of our participants reported pain during testing; however, we cannot assert if such changes in torque steadiness and coefficient of variation of motor unit discharge rate would not occur during activities that provoke pain in this group, such as running.

### 4.4 Limitations

We were unable to effectively track the same motor unit from 10 to 20% peak torque. We tried tracking the same motor units across the two intensities, but this has markedly reduced the number of motor units left for analysis. Although both intensities used in this study are considered as of low threshold, each motor unit is unique from another, and motor unit tracking would have provided more robust information about each motor unit unique response to the increase in torque. The EMG device used was limited to up to 2 × 32-channel adaptor, not allowing sampling of all three muscles at the same time. Future studies might need to consider using devices that allow recording of all three muscles during the same contractions and using electrodes with more channels (i.e. 64 electrode matrices) to increase the number of motor unit identified during decomposition when estimating neural drive to the triceps surae. Another limitation that should be highlighted is the type of contraction used for analysis. HD-EMG analysis provides reliable estimates of motor unit discharge rates^26^; however, it requires isometric contractions for motor unit analysis. Thus, the observations of neural drive from this study cannot be extrapolated into dynamic tasks such as heel raises or running. Furthermore, we used submaximal intensities of relative peak isometric torque, as this facilitates motor unit identification, therefore, it is possible that during higher torque intensities, which demand more torque of each individual muscles, the differences observed in this study would be greater.

## 5. Conclusion

Our data suggest that runners with mid-portion AT have a muscle-specific deficit in the triceps surae, possibly creating heterogeneous loading to the Achilles tendon and contributing for the high recurrencies^32^ of AT. We observed reduced motor unit discharge rate, (i.e. reduced neural drive) in GL during the increase in plantar flexor torque demands but not in GM or SOL. This deficit in motor unit excitability in GL might be greater during activities that require greater plantar flexor torque, which could contribute to overload the Achilles tendon. Different strategies to try and increase GL activation during plantar flexion resistance training could be beneficial for AT, such as adopting different feet position during heel raise. Such rehabilitation strategy should be studied in patients with AT to further understand how the reduced contribution of GL impacts Achilles tendinopathy and how implementing strategies to increase the neural drive to GL would affect AT patient outcomes.

## Data Availability

Dataset and R code are available at https://github.com/GabeFernandess/MU_TS_runners_AT

## Authors’ contribution

GLF and GST designed the study. GLF and LBRO conducted experiments. GLF analysed the data and drafted the first version of the manuscript. RLS developed a MATLAB script for motor unit discharge rate, recruitment threshold, coefficient of variation of motor unit discharge rate and torque steadiness analyses. GLF, LBRO, AS and GST critically revised the manuscript. All authors read and approved this final version of the manuscript.

## Data availability

Dataset and R code are available at https://github.com/GabeFernandess/MU_TS_runners_AT

## Acknowledgments

The authors declare having received no funding for this study.

The authors thank all the volunteers who participated in this study for their contribution to the development and achievement of this research.

## Abbreviations

AT: Achilles tendinopathy
GL: Gastrocnemius lateralis
GM: Gastrocnemius medialis
HD-EMG: High density surface electromyography
MVIC: Maximal voluntary isometric contraction
SOL: Soleus

